# Understanding the Psychological Effects of Psilocybin and 3,4-Methylenedioxymethamphetamine in a Non-Clinical Population

**DOI:** 10.1101/2025.05.28.25328532

**Authors:** Paul Bernard Fitzgerald, Samantha Lee Webb, Nigel Christopher Denning, Traill Dowie, Monica Margaret Schweickle, Anish Modak, Grace Chan, Jackie Knight, Molly Waldron, Kirsten Gainsford, Hannah Hawkes, Sonia Zammit, Novi Christina Sutanto, Bernadette Mary Fitzgibbon, Neil Wayne Bailey

## Abstract

**Objective:** Despite many decades of experimental studies and clinical trials involving a variety of psychedelic agents, we still lack a comprehensive understanding of the effects of these substances on psychological experiences. As such, we designed and conducted a study to comprehensively characterise the effects of both psilocybin and 3,4-Methylenedioxymethamphetamine (MDMA) on a range of psychological outcomes in a substantive non-clinical population.

**Methods:** This study involved a single dose administration of psilocybin or MDMA in healthy individuals in a group setting (2-4 people per session). All participants underwent a single preparation session, a drug exposure session, and an integration session within 72 hours of dosing. Outcome assessments were conducted at a pre-dosing baseline, 1-3 days post dose (side effects only), one week post dose and at 3 month follow up (the later time point data is not included here).

**Results:** Of 48 participants, 25 initially received MDMA and 23 psilocybin. Ten cross-over participants received MDMA and then psilocybin and six participants received both in the reverse order: making a total of 31 MDMA and 33 psilocybin dosing sessions. In the week after dosing, we found significant changes in personality (a reduction in neuroticism and increase in extraversion), mindfulness, and connectedness following the administration of psilocybin but not MDMA. Psilocybin also produced significantly stronger mystical experiences compared to MDMA, and there was a significant correlation between the magnitude of these mystical experiences and changes in connectedness and mindfulness (but not changes in personality). Of note, participants seemed more comfortable with, and preferred, larger group sizes when being administered MDMA than psilocybin.

**Discussion:** Our results identified a range of short-term psychological effects in non-clinical participants following a single dose of psilocybin, that were not reported following a single dose of MDMA. Notably, our results indicate that these effects following psilocybin may be moderated through its induction of mystical experiences, as has been previously hypothesised. Although preliminary, our results also suggest that larger group dosing sessions seem more feasible with MDMA than psilocybin.

## Introduction

There has been a rapid escalation of interest in the effects of a variety of psychoactive agents on human brain function and psychological experience as well as the ongoing development of therapeutic applications of agents such as psilocybin and 3,4-methylenedioxymethamphetamine (MDMA) [1]. Despite many experimental studies and clinical trials involving these drugs, we still lack a comprehensive understanding of their effects on a range of psychological outcomes which could contribute to their development as therapeutic agents [2]. As such, we designed and conducted a study to comprehensively characterise the effects of both psilocybin and MDMA on a range of aspects of neural activity, cognitive function and psychological measures in a substantive non-clinical population. This article will present data on the immediate / short term psychological changes seen with a single dose of either drug.

A range of studies have explored the short-term psychological effects of a variety of psychoactive agents (see review in [3]) including psilocybin or MDMA. However, many of these studies have examined effects only *during* the dosing session, and less research has examined changes that persist beyond this time. For example, in one of the first studies exploring the effects of psilocybin, Hassler et al examined the effects of varying doses in eight volunteers, and reported changes in consciousness, attention and physiological variables during drug exposure, but they did not study post dose effects [4]. In addition, few studies have directly compared the effects of different psychoactive agents. We note that a single study has compared the immediate effects of MDMA to other psychoactive agents (although not to psilocybin) [5], and no studies have directly compared psilocybin and MDMA to date, especially in regard to their post dose effects.

In terms of studies exploring post dose effects, one of the main areas of focus has been on examining the impact on these drugs on mood and empathy. Short term effects of psilocybin on mood and empathy related variables have been reported in healthy control groups across a small series of studies [6]. For example, in one study positive affect remained increased one month post dose in a small group of healthy subjects, although initially reported reductions in negative affect had subsided [7]. One recent larger study found a short-term trend for an increase in mean negative affect scores on the PANAS following a 25mg psilocybin dose [8]. Studies have also found short term increases in emotional empathy and well-being when psilocybin was administered in a social setting [9], as well as increases in emotional but not cognitive empathy [10].

In addition to the effects on mood and empathy, it has been proposed that one of the mechanisms of therapeutic change with these drugs may be through modification of aspects of psychological function classically described as domains of personality, especially aspects such as openness as typically assessed in five factor personality models [11]. Effects of psilocybin on measures of personality have been studied in both healthy volunteers and in patients with depression. In healthy individuals, increases in openness have been found to persist over time following dosing [12] and in clinical populations, decreased neuroticism, increased extraversion, and increased openness have been reported [13].

In addition to changes in personality, the nature and magnitude of mystical experiences that occur during medication dosing have been proposed as a significant predictor of possible therapeutic effects arising from psychedelic medication use in previous research [14]. The Mystical Experiences Questionnaire (MEQ) [15] has been used to quantify these, with more mystical experiences seemingly predictive of longer term positive emotional outcomes following dosing in healthy volunteers [16]. A second line of enquiry has examined the impact of these medications, especially psilocybin, on trait mindfulness, with some suggestion of positive effects [17].

Although a wide range of within dosing session effects of MDMA have been reported including on mood and somatic experiences [18], few studies have explored post dose effects and much of this research has focused on recreational use and regular users. Studies of the effects of MDMA on mood in clinical studies have failed to find the post dose decline in mood that has been reported to be associated with recreational consumption [19]. Studies in controlled conditions have not consistently shown the types of pro-social and empathy related effects reported in other types of studies [20]. For example, a controlled study that administered a single 100mg dose of MDMA to healthy participants then analysed post dose effects at 3 days follow up showed reduced anxiety, but no changes in mood, trust, pro-social inclinations or empathy [20]. However, this study, along with others that have reported positive effects of MDMA on aspects of empathy and social behaviour [21,22], was conducted in MDMA or polydrug using subjects.

In summary, while there is emerging evidence supporting the therapeutic value of psilocybin therapy in depression and MDMA in the treatment of post-traumatic stress disorder (PTSD), we lack a comprehensive understanding of the effects of these drugs. Therefore, we undertook a study to characterise the effects of both psilocybin and MDMA in healthy, currently non-drug using, subjects on a variety of psychological, cognitive and physiological / neural measures. The aim of this article is to describe the psychological effects across domains of mood and anxiety, personality, trust, mindfulness, connectedness and mystical experiences seen in the week post dosing. We aimed to explore whether there were differences between the mystical experiences reported by subjects using the two different drugs and whether the mystical experiences reported were related to any emergent changes on the outcome variables. In this regard we hypothesised that psilocybin would produce a greater range of mystical experiences, and these would be more strongly correlated with changes in personality, trust, mindfulness, metaphysical beliefs and connectedness.

## Methods

### Study Design

The primary study (“A study of the psychological, cognitive and physiological effects of Psychedelic Medicines” [the ASSESS trial]) involved a comparison between the effects of a single dose of psilocybin and MDMA, evaluating the persistence of psychological effects of drug exposure one week and 3 months after drug exposure. The study was conducted in accordance with the Declaration of Helsinki and International Conference on Harmonization Guidelines in Good Clinical Practice and the study was approved by the Human research Ethics Committees of ACT Health and the Australian National University. All subjects provided informed consent and the study was registered with Australian New Zealand Clinical Trials Registry (ACTRN12622001535763p, https://anzctr.org.au/ACTRN12622001535763.aspx, date of registration: 12/12/2022).

### Participants

Initially, recruitment of 100 participants was planned, but recruitment was abbreviated due to resource issues. Recruitment and assessments took place from 2023 to 2024. A total of 50 healthy individuals consented to participate. Two participants withdrew prior to medication dosing, leaving a total of 48 participants: 14 male (age 47.8±13.2) and 34 female (age 49.8±8.8). Participants could choose the drug they were to take in the dosing session and there was an option to participate twice, taking the alternative drug, after a minimum interval between doses of three months. All participants within each session took the same drug.

Of the 48 participants, 25 initially received MDMA and 23 psilocybin. Ten participants received MDMA and then psilocybin and six participants received both in the reverse order. Therefore, there were a total of 31 MDMA and 33 psilocybin doses administered. For the MDMA sessions, ten participants participated in dosing sessions with group sizes of 2, six in groups of 3 and 15 participants completed the dosing sessions in groups of 4. For psilocybin, there were six participants in groups of 2, 15 participants in groups of 3, and 12 participants in groups of 4. In the MDMA sessions, the supplementary additional half dose was provided to 26 of the 31 participants.

All participants were individuals with interest in psychedelic assisted psychotherapy (12 psychiatrists or other doctors, 21 psychologists, 10 counsellors or psychotherapists, and 5 others (nursing, research backgrounds)) who had undertaken training in psychedelic assisted psychotherapy through several local training providers. This provided an opportunity for therapists in training to have an experience of psychedelic medication sessions. Due to their previous education in psychedelic therapies, participants had an awareness of what to expect in a dosing session, including expected effects and side effects of each drug.

All participants were between 18 and 70 years of age, able to provide informed consent, had a support person available for the night after the dosing session and were proficient in written and spoken English. Exclusion criteria included pregnancy, unstable medical illness, a form of cardiovascular disease that could pose a risk to the participant during the dosing session, taking psychoactive medications, a history of seizures or moderate to severe traumatic brain injury, hepatic impairment, insulin-dependent diabetes, a current or previous diagnosis of schizophrenia spectrum, other psychotic disorders, bipolar disorder, alcohol or other substance dependence. Participants were excluded if they had used either MDMA or psilocybin in the past 3 months or substantially in the past (defined as more than 100 drug exposures).

### Study Procedures

Following informed consent and assessment for study suitability, the process of the study for each individual involved a single preparation session where a study therapist briefed participants on what to expect in the dosing session, the drug exposure session, and an integration session within 72 hours of dosing. Assessments were conducted at a pre-dosing baseline (within one week of the dosing session), 1-3 days post dose (side effects only), one week post dose (window 5-16 days) and at a 3 month follow up timepoint.

The preparation sessions were conducted by an experienced therapist online or in person and included all members of the group. Participants were provided with written documents outlining the process of the dosing sessions and expectations of them before and during the session. Dosing sessions were conducted in a large room designed to provide a low stimulus and calm environment with calm curated music playing, set in a mental health clinic in Melbourne, Australia. The sessions were conducted in groups of 2-4 individuals, monitored by two experienced therapists and with available medical support. Participants could sit and engage in conversation or lie quietly using eye shades. A second room was available, so that participants who were undergoing distress of any form could be moved to that room for increased attention from one of the two therapists without disrupting the other participants. Blood pressure, heart rate and body temperature were measured prior to dosing and once per hour up to the first 3 hours after dosing and prior to discharge by a trained research team member. Participants completed a side effects questionnaire before leaving the session in the company of a support person.

Within 72 hours of the psychedelic exposure session, participants undertook an integration session online and included all members of the group. Participants discussed the experiences they had during the session and the nature of their reactions during the sessions and afterwards.

### Psilocybin and MDMA

For the psilocybin arm of the study, a single dose in capsule form of 25 mg of psilocybin was provided for a person weighing under 90 kg or 30 mg for a person weighing between 90 kg and 115Kg (there were no participants above this weight). All subjects who received MDMA received an initial 80mg dose with a supplementary dose of 40mg available if there were inadequate effects after 1 hour (at the discretion of the lead therapist, participant, and study doctor).

Both drugs were supplied through a contractual relationship with Mind Medicine Australia (MMA) who sourced supply of psilocybin from Psygen Labs Inc. and MDMA from Dalton Chemical Laboratories Inc as well as OPTIMI Labs Inc. for supply of both Psilocybin and MDMA. All medications supplied were medical grade Good Manufacturing Practice (GMP) standard, with a Certificate of Analysis and GMP certification. Both medications were imported by a licenced pharmacy and stored under the conditions determined in the appropriate government importing and storage permits. Drug doses were couriered to the study site after trial medication prescription and the granting of an individual participant permit through the local health department processes.

### Study Measures

A series of measures were completed at baseline, one week and 3-month post dose. These included the Depression and anxiety stress scale (DASS) [23] (this is a 40 item self-report measure of depression, anxiety and stress), the State and Trait Anxiety Index (STAI) [24] (a commonly used validated 20 item measure of trait and state anxiety), the IPIP-NEO-60 [25] (a brief (60 item) five factor model-based personality inventory based on the International Personality Item Pool (IPIP)), the General Trust Scale (GTS) [26] (a self-reported scale measuring trust in other people), the Mystical Experience Questionnaire (MEQ) [15] (assessing mystical experiences across 4 dimensions: mystical (including items from the internal unity, external unity, noetic quality), positive mood, transcendence of time and space, and ineffability), the Mindful Attention Awareness Scale (MAAS) [27] (assessing individual differences in the frequency of mindfulness states in day-to-day life, defined as attention to and awareness of the experience of the present moment), the Metaphysical Beliefs Questionnaire (MBQ) [28] (assessing metaphysical beliefs related to ones understanding of mental and physical reality generating a ‘non-physicalist belief’ score and a ‘physicalist belief’ score), and the Watts Connectedness Scale (WCS) [29] (assessing the participant’s sense of their connection to self, others and the world in three subscale scores).

The effects of group size were assessed via two questions measured at 1 week follow up and 3 month follow up (only data from the 1 week follow up is presented in this paper). First, participants were asked “Did you feel comfortable with the number of people in your session?” with a response selected from “0=not comfortable to 7=completely comfortable”. They were then asked “Would you have preferred?”, with the following options: Less people / More people / It felt like the right amount of people”.

Participants also underwent EEG and cognitive assessments at baseline and the one-week post dose assessment and self-report rating of their confidence as psychedelic assisted psychotherapists. This data and data from the STAI will be reported separately. Detailed data rated on a 6-point scale was collected on the experience of thirty possible side effects at the end of the session, the following day, after 3 and 7 days at after 3 months. This side effect data will also be reported separately.

### Data Analysis

Paired samples t-tests were used to analyse whether there were changes in a series of outcome measures from baseline to the post dosing one week follow up assessment time point for the two medication groups separately, including data from all dosing sessions. Repeated measures ANOVA models were used to explore differences in pre- to post changes between the groups on measures of mood and anxiety from the DASS. Independent samples t-tests were used to compare the Mystical Experience Questionnaire data between the 2 medication groups. Pearson correlations were calculated between the Mystical Experience Questionnaire subscale scores and any outcomes measures where changes were seen from baseline to 1 week follow up. ANOVA models were used to explore the effects of group size. All analysis was conducted in SPSS 29.0 (2022).

## Results

### DASS Scores

DASS scores before and after dosing for the 2 medication groups are presented in Table 1. All baseline scores were in the normal range. The mean values for total, anxiety and depression scores reduced across time in both medication groups: the reductions in Total and Anxiety scores approached significance in the psilocybin group. The repeated measures ANOVA analysis assessing change over time between the 2 groups showed effects of time (an overall reduction) for Total DASS scores (p=0.03) and Anxiety (p=0.04) but not for Depression (p=0.52). There were no differences between the 2 drug groups (TIME X DRUG interactions: Total p=0.89, Anxiety p=0.90, Depression p=0.93).

**Table 1:** DASS scores from baseline to post dosing follow up.

|  |  | Baseline |  | Post Dose |  |  |
| --- | --- | --- | --- | --- | --- | --- |
|  |  | Mean | SD | Mean | Sd | P |
| MDMA | Total | 2.6 | 2.1 | 2.0 | 2.5 | 0.20 |
|  | Anxiety | 1.9 | 1.4 | 1.4 | 2.3 | 0.25 |
|  | Depression | 0.8 | 1.1 | 0.7 | 1.0 | 0.43 |
| Psilocybin | Total | 3.1 | 2.0 | 2.4 | 2.4 | 0.08 |
|  | Anxiety | 2.0 | 1.3 | 1.5 | 1.5 | 0.07 |
|  | Depression | 1.1 | 1.3 | 1.0 | 1.7 | 0.58 |

### IPIP-NEO Personality Dimensions

IPIP scores before and after dosing for the 2 medication groups are presented in Table 2. There were no significant changes after MDMA sessions, but the psilocybin sessions resulted in a reduction in neuroticism (p=0.03) and increase in extraversion (p=0.01).

**Table 2:** IPIP – NEO scores from baseline to post dosing follow up.

|  |  | Baseline |  | Post Dose |  |  |
| --- | --- | --- | --- | --- | --- | --- |
|  |  | Mean | SD | Mean | Sd | P |
| MDMA | Neuroticism | 24.9 | 7.5 | 23.7 | 7.8 | 0.28 |
|  | Extraversion | 46.5 | 8.4 | 45.4 | 7.8 | 0.14 |
|  | Openness | 47.0 | 6.6 | 47.3 | 7.0 | 0.53 |
|  | Agreeableness | 52.1 | 5.3 | 51.8 | 5.6 | 0.45 |
|  | Conscientiousness | 48.5 | 6.4 | 49.1 | 6.0 | 0.36 |
| Psilocybin | Neuroticism | 23.6 | 7.4 | 21.8 | 7.7 | 0.03 |
|  | Extraversion | 43.6 | 6.9 | 45.1 | 6.1 | 0.01 |
|  | Openness | 48.4 | 6.1 | 48.0 | 6.1 | 0.40 |
|  | Agreeableness | 52.6 | 4.7 | 52.9 | 4.8 | 0.58 |
|  | Conscientiousness | 49.1 | 5.9 | 49.6 | 5.9 | 0.38 |

### Trust in Others, Connectedness, Beliefs and Mindfulness

GTS scores before and after dosing for the 2 medication groups are presented in Table 3. There was no change in trust in either group. For the WCS, there was no changes in connectedness observed in the MDMA condition but in the psilocybin condition, there were increases in connectedness to oneself (p=0.01), the world (p=0.008) and in general (p=0.007).

**Table 3:** General Trust Scale and Watts Connectedness Scale scores from baseline to post dosing follow up.

|  |  |  | Baseline |  | Post Dose |  |  |
| --- | --- | --- | --- | --- | --- | --- | --- |
|  |  |  | Mean | SD | Mean | Sd | P |
| GTS | MDMA |  | 5.52 | 0.93 | 5.49 | 1.0 | 0.85 |
|  | Psilocybin |  | 5.50 | 0.88 | 5.41 | 1.0 | 0.42 |
| WCS | MDMA | To self | 80.11 | 14.46 | 82.22 | 13.71 | 0.33 |
|  |  | To others | 79.56 | 16.19 | 79.54 | 15.40 | 0.99 |
|  |  | To world | 70.00 | 20.38 | 73.79 | 20.28 | 0.18 |
|  |  | General | 76.56 | 14.69 | 78.51 | 14.08 | 0.31 |
|  | Psilocybin | To self | 76.29 | 13.98 | 81.88 | 12.46 | **0.01** |
|  |  | To others | 73.67 | 15.74 | 77.67 | 17.40 | 0.15 |
|  |  | To world | 65.29 | 19.78 | 75.64 | 17.65 | **<0.01** |
|  |  | General | 71.75 | 13.32 | 78.40 | 13.37 | **<0.01** |

MBQ scores are presented in Table 4: there were no changes in Physicalist or Non-Physicalist beliefs in either medication condition.

**Table 4:**
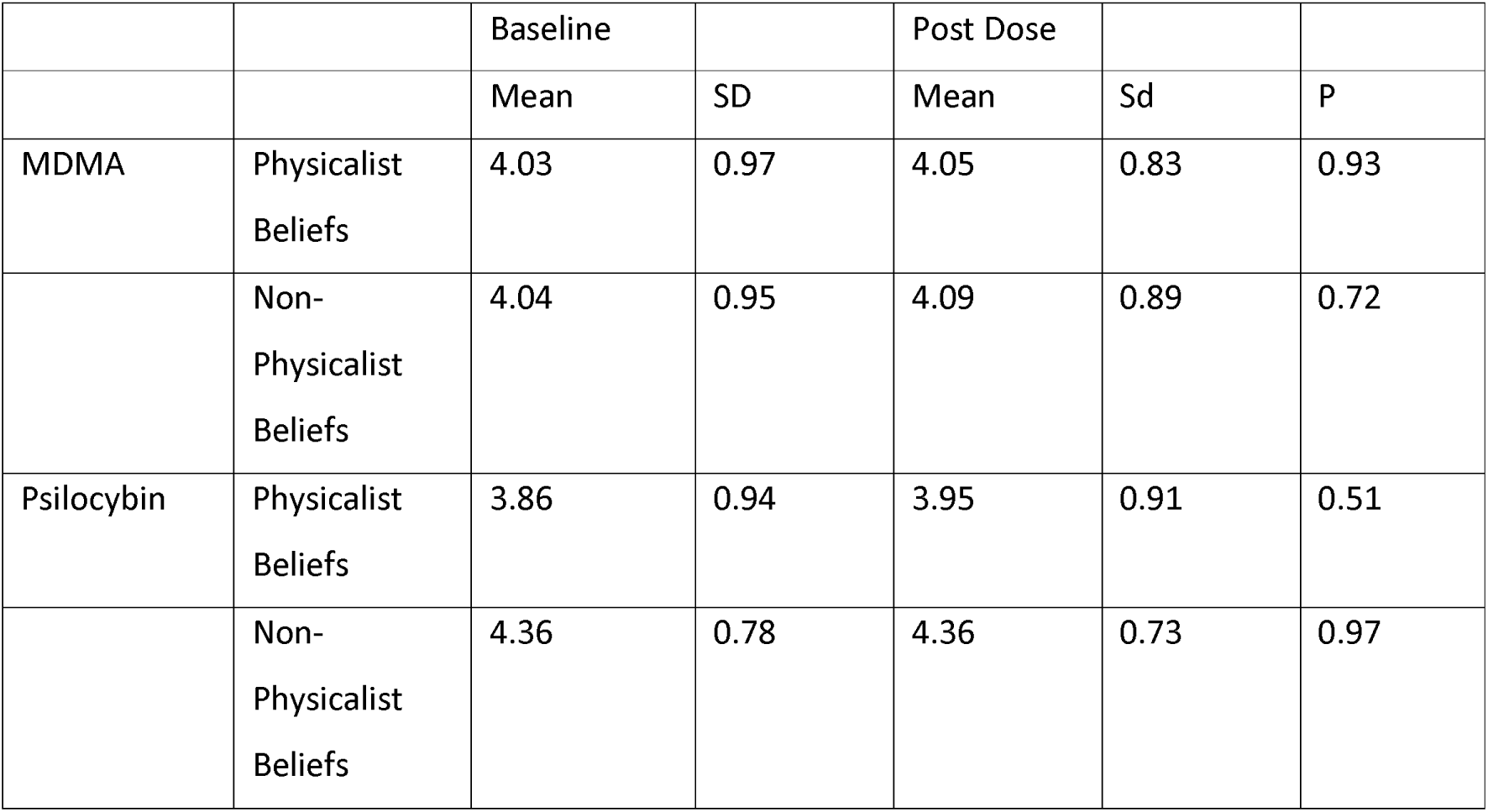
Metaphysical Beliefs Questionnaire from baseline to post dosing follow up.

Mindful Attention Awareness Scale scores are presented in Table 5. Psilocybin produced a significant increase in dispositional mindfulness (p=0.003), but MDMA did not (p=0.62).

**Table 5:**
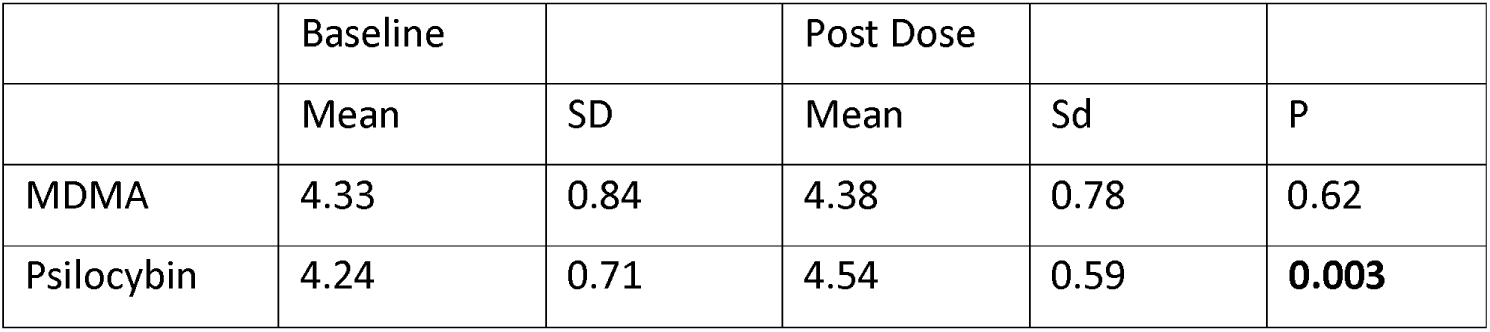
Mindful Attention Awareness Scale from baseline to post dosing follow up.

### Mystical Experiences

There were significant differences between conditions in the mystical experiences reported by participants. Specifically, participants reported that they experienced a greater sense of transcendence (p=0.001), ineffability (p=0.016) and overall / average scores (p=0.045) when they had received psilocybin compared to when they had received MDMA. There were no differences in positive mood (p=0.686) or mystical experiences as a subscale (p=0.091).

The magnitude of mystical experiences in all dimensions were correlated with the increase in total connectedness found on the WCS following psilocybin dosing (all r > 0.35, all p < 0.04), as well as self-connectedness (all r > 0.45, all p < 0.01). The average mystical experience score, as well as the mystical score and positive mood dimensions also correlated with the world connectedness dimension of the WCS (both r > 0.39, both p < 0.025). The average, mystical, and transcendence dimensions of the mystical experience scale also correlated with increases in mindfulness (all r > 0.35, all p < 0.05, Table 6). There were no relationships between the magnitude of mystical experiences and changes in neuroticism and extraversion.

**Table 6:** The relationship between mystical experiences and changes in connectedness, mindfulness, neuroticism and extraversion.

|  |  |  | WCS |  |  | MAAS | IPIP-NEO |  |
| --- | --- | --- | --- | --- | --- | --- | --- | --- |
|  |  |  | Total | World | Self |  | Neuroticism | Extraversion |
| MEQ | Average Score | Correlation | **0.484** | **0.428** | **0.526** | **0.385** | -0.028 | 0.075 |
|  |  | Sig | **0.004** | **0.013** | **0.002** | **0.027** | 0.875 | 0.676 |
|  | Mystical Score | Correlation | **0.434** | **0.395** | **0.472** | **0.357** | 0.000 | 0.050 |
|  |  | Sig | **0.012** | **0.023** | **0.006** | **0.042** | 1.000 | 0.784 |
|  | Positive Mood | Correlation | **0.526** | **0.487** | **0.486** | 0.322 | 0.037 | 0.130 |
|  |  | Sig | **0.002** | **0.004** | **0.004** | 0.067 | 0.837 | 0.470 |
|  | Transcendence | Correlation | **0.417** | **0.307** | **0.513** | **0.412** | -0.126 | 0.117 |
|  |  | Sig | **0.016** | **0.082** | **0.002** | **0.017** | 0.483 | 0.517 |
|  | Ineffability | Correlation | **0.367** | **0.350** | **0.455** | 0.233 | -0.118 | -0.063 |
|  |  | Sig | **0.036** | **0.046** | **0.008** | 0.192 | 0.514 | 0.728 |

### Tolerability

There were no serious adverse events. Reported adverse events are presented in Table 7. Several participants experienced some degree of ongoing distress after dosing and required additional integration sessions. Five participants had elevated blood pressure at the end of the dosing days (at discharge (2 psilocybin, 3 MDMA). All returned to the appropriate range or were deemed safe to leave by the study doctor on day of dosing. One participant was requested to repeat their blood pressure reading the following day which returned to the normal range.

**Table 7.** Reported adverse events (each reported in a single participant)

|  |  |
| --- | --- |
| Psilocybin |  |
| Severe headache | Dosing to day 2 post dose, fully resolved |
| Pain in side of abdomen during dosing | Resolved with ingestion of food |
| Persistent distress post dose | Resolved with additional integration sessions |
| Persistent distress post dose | Lessened with additional integration sessions |
| Feeling flat, apathetic, meaningless post dose | Resolved with additional integration session |
| MDMA |  |
| Fatigue/muscle weakness/lack of appetite | Persisted for 2 weeks post dose, resolved following additional integration session |

Overall, there were fewer side effects reported with MDMA than psilocybin, although this was only significant for the dosing day. The sum of all side effects reported to be experienced by an individual on the dosing day for MDMA was 24.3±9.3 and for psilocybin was 41.4±13.8 (p<0.001). One day post-dosing session, these values were 17.7±12.0 for MDMA and 10.4±11.4 for psilocybin (p=0.44). Three days after the dosing session, these values were 8.7±9.4 for MDMA and 10.4±11.4 for psilocybin (p=0.27). Seven days later, these values were 9.6±12.3 for MDMA and 9.2±9.3 for psilocybin (p=0.90).

### Group Effects

Dosing sessions were conducted in groups ranging from 2 to 4 participants (Figure 1). At the 1 week post dosing timepoint, for MDMA, there was no difference in the overall reported comfort that individuals felt during their dosing session based on group size (F(2,28) = 0.7, p = 0.93), and there were no differences in reported group size preference (F(2,28) = 0.31, p = 0.74). As shown in Figure 1, most subjects reported a preference for the group size they had experienced. Three (of 10) people who participated in dosing session groups of 2 reported preferring fewer people in the session. One of six participants in the groups of 3 reported preferring fewer people in the session, and four people (of 15) who participated in dosing sessions in groups of 4 reported preferring fewer people in the session.

**Figure 1:**
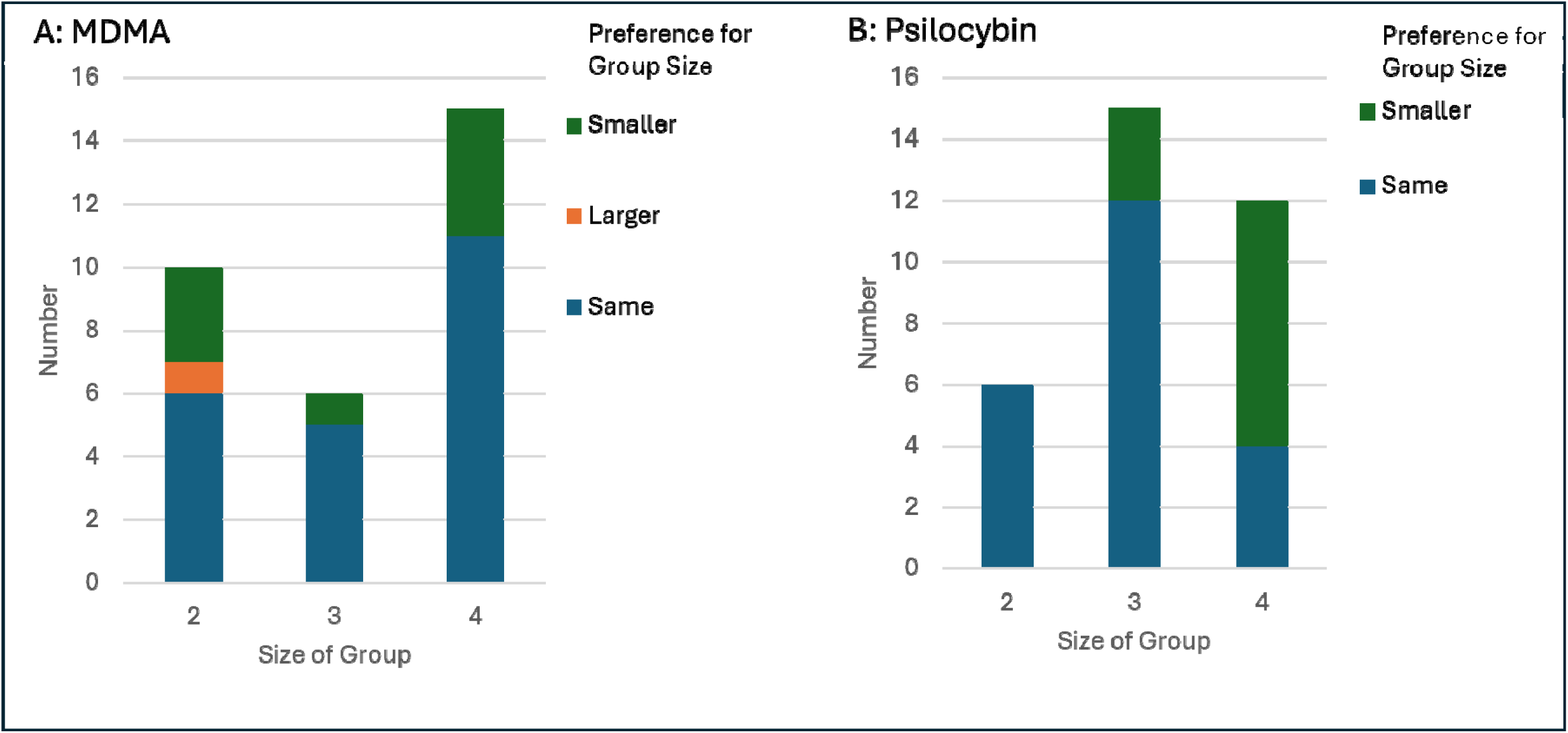
Preference for group size: A. for MDMA, B. for Psilocybin

At the 1 week post dosing timepoint, for psilocybin, there was an overall difference between group sizes on reported group comfort (F(2,30) = 3.4, p = 0.045). There was greater reported group size satisfaction in those who participated in dosing session groups of 2 versus those in groups of 4 (6.8±0.41 versus 4.8±2.1, p=0.024) but no significant difference between the group sizes of 2 and 3, and no significant difference between group sizes of 3 and 4.

There was also a significant difference in group size preference in the psilocybin condition (F(2,30) = 6.7, p = 0.004). All of the subjects in group sizes of 2 reported a preference for their group size. Three (of 15) participants in group sizes of 3 and a majority (8 of 12) participants in group sizes of 4 preferred a smaller group.

## Discussion

In this study we explored the effects of psilocybin and MDMA on a variety of aspects of psychological experiences when the medications were provided to healthy controls in a supported dosing session similar to those used during the provision of psychedelic assisted psychotherapy. We found that in the week after dosing, there were significant changes in personality, mindfulness, and connectedness following the administration of psilocybin but not MDMA. Specifically, psilocybin produced a reduction in neuroticism and an increase in extraversion on a brief five factor model assessment of personality, it enhanced the sense of individuals’ connectedness to themselves and to the world around them and increased their self-reported dispositional mindfulness. Psilocybin produced significantly greater mystical experiences compared to MDMA, and there was a significant correlation between the magnitude of these mystical experiences and changes in connectedness and mindfulness (but not changes in neuroticism and extraversion). Of note, participants seemed more comfortable with larger group sizes when being administered MDMA than with psilocybin, with most participants in the MDMA sessions reporting a preference for the group size they participated in, and no differences in satisfaction with the group size reported between the different group sizes. In contrast, most participants in groups of four participants in the psilocybin condition reported they would have preferred a smaller group size, and a significant difference was present between group size satisfaction ratings for the group sizes of four compared to group sizes of two.

Previous research has reported some overlapping findings in regard to the effects of psilocybin, and consistent with our results there is much less evidence of an impact of MDMA across the studied domains. Regarding the aspects of personality assessed with the IPIP-NEO, there is some divergence between our results and the previous literature. Specifically, we found significant changes in extraversion and neuroticism which have been reported in clinical (depressed) but not non-clinical populations [13]. In contrast, research in non-clinical populations has reported increased openness [12] which we did not find. Interestingly, changes in openness in a previous study were associated with the self-reported magnitude of within session mystical experiences [12]. A similar relationship was shown between changes in personality variables and ‘insightfulness’ experienced during medication dosing [13] assessed with the altered state of consciousness questionnaire (ASC) [30].

Our results suggest the possibility that the impacts of psilocybin are moderated through its induction of mystical experiences, as has been previously hypothesised [31]. Consistent with the effects of psilocybin (as a more traditional psychedelic drug, compared to MDMA), participants who received psilocybin reported substantially greater mystical experiences across several domains compared to MDMA. It is notable that there were significant correlations between the mystical experiences of participants who had received psilocybin and changes in connectedness and mindfulness. This suggests a potential mechanistic pathway for the therapeutic effects of psilocybin, where mystical experiences during the dosing session are associated with lasting increases in feelings of connectedness and in dispositional mindfulness, which previous (non-psychedelic) research has indicated are associated with better mental health [32]. However, perhaps somewhat puzzling, mystical experiences within the dosing session were not associated with changes in the personality variables of neuroticism and extraversion. It may be that those personality factors are modified via a different pathway (that our measures did not capture) compared to the changes in feelings of connectedness and dispositional mindfulness.

Furthermore, it is interesting that MDMA was not associated with an increase in feelings of trust or connectedness. This may suggest that the prosocial and empathy effects of MDMA are restricted to the acute period of administration, and do not elicit lasting changes. However, this tentative conclusion must be caveated with the awareness that our sample was restricted to healthy individuals. Recent evidence suggests MDMA may be effective as a treatment for PTSD and social anxiety disorder, with increased trust and therapeutic alliance suggested to be part of the mechanism of action [33–35]. While our results do not indicate MDMA exposure was associated with effects on trust or connectedness measured outside of the dosing session, it may be that our healthy participants were at a ceiling level on these measures, with less potential range for change from before to after the MDMA session, and that clinical populations with potentially lower levels of these variables at baseline may show larger changes. This may be even more the case in our sample which consisted of health care professionals who may be more trusting and connected than average members of the community.

In the context of the lack of effects from MDMA and a wide range of effects from psilocybin, we note that both substances were administered within the range of typical clinical doses. However, despite the real-world applicability of our findings, from a purely physiological perspective, it may be that the psilocybin dose produced stronger effects because the psilocybin dose was relatively higher / more psychoactive than the MDMA dose. Clinical doses of MDMA are more limited by potential risk factors than psilocybin doses, and from a purely theoretical perspective, our conclusions are necessarily limited by an inability to determine if the doses were matched. It may be that MDMA would show stronger effects at higher doses, but testing this idea is not safe in human participants. Additional limitations of our study are the lack of a control condition (which means we cannot draw causal inferences), a relatively small sample size for each condition, and restriction of our sample to participants who have received training in psychedelic assisted psychotherapy (potentially limited the generalisability of our results).

In conclusion, we found significant effects of psilocybin across a range of measures of psychological functioning, following a single dose in a well-informed group of non-clinical subjects. However, while the pre- to post-dosing effects that we detected for the psilocybin condition were spread across a large range of psychological factors, we did not detect any significant effects for the MDMA condition. Effects of psilocybin on connectedness and mindfulness, but not changes in neuroticism and extraversion, were related to the degree of mystical experiences reported following dosing sessions. Larger group dosing sessions seem more feasible with MDMA than psilocybin based on group preferences and reported group size comfort.

## Ethics Information

Ethics approval was provided by the ACT Health Human Ethics Committee, with reciprocal approval from the Australian National University Human Ethics Committee. All participants provided written informed consent prior to participation in the study.

## Data Availability Statement

The de-identified data that support the findings of this study are available from the corresponding author, PBF, upon reasonable request.

## References

1 O’Donnell KC, Grigsby J, Grob CS. Healing, Harms, and Humility: Expanding the Scope of Psychedelic-Assisted Psychotherapy Research. Am J Psychiatry. 2025;182(1):13–16.

2 Miceli McMillan R, Fernandez AV. Understanding subjective experience in psychedelic-assisted psychotherapy: The need for phenomenology. Aust N Z J Psychiatry. 2023;57(6):783–88.

3 Evens R, Schmidt ME, Majic T, Schmidt TT. The psychedelic afterglow phenomenon: a systematic review of subacute effects of classic serotonergic psychedelics. Ther Adv Psychopharmacol. 2023;13:20451253231172254.

4 Hasler F, Grimberg U, Benz MA, Huber T, Vollenweider FX. Acute psychological and physiological effects of psilocybin in healthy humans: a double-blind, placebo-controlled dose-effect study. Psychopharmacology (Berl). 2004;172(2):145–56.

5 Holze F, Vizeli P, Muller F, Ley L, Duerig R, Varghese N, et al. Distinct acute effects of LSD, MDMA, and D-amphetamine in healthy subjects. Neuropsychopharmacology. 2020;45(3):462–71.

6 Heuschkel K, Kuypers KPC. Depression, Mindfulness, and Psilocybin: Possible Complementary Effects of Mindfulness Meditation and Psilocybin in the Treatment of Depression. A Review. Front Psychiatry. 2020;11:224.

7 Barrett FS, Doss MK, Sepeda ND, Pekar JJ, Griffiths RR. Emotions and brain function are altered up to one month after a single high dose of psilocybin. Sci Rep. 2020;10(1):2214.

8 Rucker JJ, Marwood L, Ajantaival RJ, Bird C, Eriksson H, Harrison J, et al. The effects of psilocybin on cognitive and emotional functions in healthy participants: Results from a phase 1, randomised, placebo-controlled trial involving simultaneous psilocybin administration and preparation. J Psychopharmacol. 2022;36(1):114–25.

9 Mason NL, Mischler E, Uthaug MV, Kuypers KPC. Sub-Acute Effects of Psilocybin on Empathy, Creative Thinking, and Subjective Well-Being. J Psychoactive Drugs. 2019;51(2):123–34.

10 Pokorny T, Preller KH, Kometer M, Dziobek I, Vollenweider FX. Effect of Psilocybin on Empathy and Moral Decision-Making. Int J Neuropsychopharmacol. 2017;20(9):747–57.

11 McCrae RR, John OP. An introduction to the five-factor model and its applications. J Pers. 1992;60(2):175–215.

12 MacLean KA, Johnson MW, Griffiths RR. Mystical experiences occasioned by the hallucinogen psilocybin lead to increases in the personality domain of openness. J Psychopharmacol. 2011;25(11):1453–61.

13 Erritzoe D, Roseman L, Nour MM, MacLean K, Kaelen M, Nutt DJ, et al. Effects of psilocybin therapy on personality structure. Acta Psychiatr Scand. 2018;138(5):368–78.

14 Reiff CM, Richman EE, Nemeroff CB, Carpenter LL, Widge AS, Rodriguez CI, et al. Psychedelics and psychedelic-assisted psychotherapy. American Journal of Psychiatry. 2020;177(5):391–410.

15 Barrett FS, Johnson MW, Griffiths RR. Validation of the revised Mystical Experience Questionnaire in experimental sessions with psilocybin. Journal of Psychopharmacology. 2015;29(11):1182–90.

16 McCulloch DE, Grzywacz MZ, Madsen MK, Jensen PS, Ozenne B, Armand S, et al. Psilocybin-Induced Mystical-Type Experiences are Related to Persisting Positive Effects: A Quantitative and Qualitative Report. Front Pharmacol. 2022;13:841648.

17 Madsen MK, Fisher PM, Stenbæk DS, Kristiansen S, Burmester D, Lehel S, et al. A single psilocybin dose is associated with long-term increased mindfulness, preceded by a proportional change in neocortical 5-HT2A receptor binding. European Neuropsychopharmacology. 2020;33:71–80.

18 Baylen CA, Rosenberg H. A review of the acute subjective effects of MDMA/ecstasy. Addiction. 2006;101(7):933–47.

19 Sessa B, Aday JS, O’Brien S, Curran HV, Measham F, Higbed L, et al. Debunking the myth of ‘Blue Mondays’: No evidence of affect drop after taking clinical MDMA. J Psychopharmacol. 2022;36(3):360–67.

20 Borissova A, Ferguson B, Wall MB, Morgan CJ, Carhart-Harris RL, Bolstridge M, et al. Acute effects of MDMA on trust, cooperative behaviour and empathy: A double-blind, placebo-controlled experiment. J Psychopharmacol. 2021;35(5):547–55.

21 Kirkpatrick MG, Lee R, Wardle MC, Jacob S, de Wit H. Effects of MDMA and Intranasal oxytocin on social and emotional processing. Neuropsychopharmacology. 2014;39(7):1654–63.

22 Kuypers KP, de la Torre R, Farre M, Yubero-Lahoz S, Dziobek I, Van den Bos W, et al. No evidence that MDMA-induced enhancement of emotional empathy is related to peripheral oxytocin levels or 5-HT1a receptor activation. PLoS One. 2014;9(6):e100719.

23 Lovibond PF, Lovibond SH. The structure of negative emotional states: Comparison of the Depression Anxiety Stress Scales (DASS) with the Beck Depression and Anxiety Inventories. Behaviour Research and Therapy. 1995;33(3):335–43.

24 Finch AJ, Kendall PC, Newmark CS, Faschiningbauer TR. Factor analysis of the MMPI-STAI. J Clin Psychol. 1975;31(3):449–52.

25 Maples-Keller JL, Williamson RL, Sleep CE, Carter NT, Campbell WK, Miller JD. Using Item Response Theory to Develop a 60-Item Representation of the NEO PI–R Using the International Personality Item Pool: Development of the IPIP–NEO–60. Journal of Personality Assessment. 2019;101(1):4–15.

26 Horii T, Tsuchiya E. A study of the relationship between the earliest recollection and interpersonal trust. The Japanese Journal of Personality. 1995;3:27–36.

27 Brown KW, Ryan RM. The benefits of being present: mindfulness and its role in psychological well-being. Journal of Personality and Social Psychology. 2003;84(4):822.

28 Timmermann C, Kettner H, Letheby C, Roseman L, Rosas F, Carhart-Harris R. Psychedelics alter metaphysical beliefs. 2021.

29 Watts R, Kettner H, Geerts D, Gandy S, Kartner L, Mertens L, et al. The Watts Connectedness Scale: a new scale for measuring a sense of connectedness to self, others, and world. Psychopharmacology (Berl). 2022;239(11):3461–83.

30 Dittrich A. The standardized psychometric assessment of altered states of consciousness (ASCs) in humans. Pharmacopsychiatry. 1998;31 Suppl 2:80–4.

31 Griffiths R, Richards W, Johnson M, McCann U, Jesse R. Mystical-type experiences occasioned by psilocybin mediate the attribution of personal meaning and spiritual significance 14 months later. J Psychopharmacol. 2008;22(6):621–32.

32 Tomlinson ER, Yousaf O, Vitterso AD, Jones L. Dispositional Mindfulness and Psychological Health: a Systematic Review. Mindfulness (N Y). 2018;9(1):23–43.

33 Johansen PO, Krebs TS. How could MDMA (ecstasy) help anxiety disorders? A neurobiological rationale. J Psychopharmacol. 2009;23(4):389–91.

34 Luoma JB, Shahar B, Kati Lear M, Pilecki B, Wagner A. Potential processes of change in MDMA-Assisted therapy for social anxiety disorder: Enhanced memory reconsolidation, self-transcendence, and therapeutic relationships. Hum Psychopharmacol. 2022;37(3):e2824.

35 Zeifman RJ, Kettner H, Ross S, Weiss B, Mithoefer MC, Mithoefer AT, et al. Preliminary evidence for the importance of therapeutic alliance in MDMA-assisted psychotherapy for posttraumatic stress disorder. Eur J Psychotraumatol. 2024;15(1):2297536.

